## Supplementary Information for "BharatSim: An agent-based modelling framework for India"

October 13, 2024

#### S1 Appendix: Scaling BharatSim with population size and model complexity

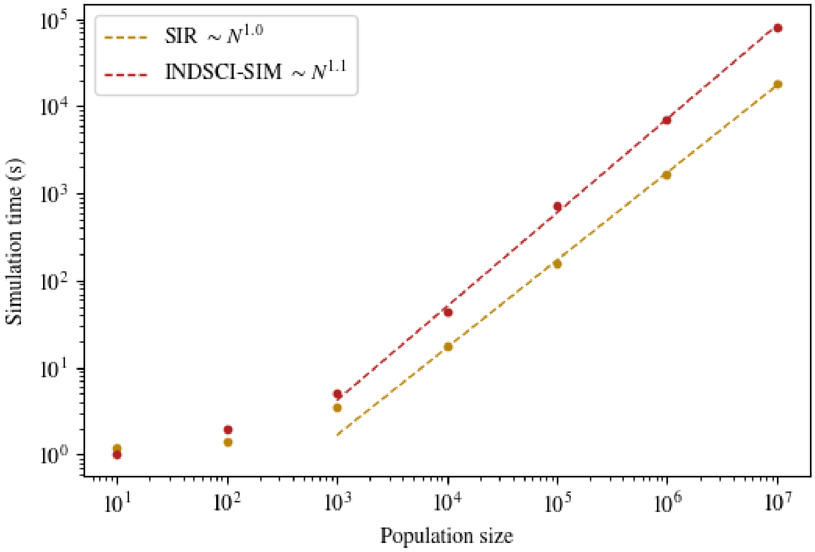


**Fig S1.1: Scaling up BharatSim.** The times taken to simulate two different models are plotted as a function of population size. In the first, we consider a simple SIR model, and in the second we consider the more complex 9-compartment INDSCI-SIM model that this paper is based on. These times are found to scale roughly as $\mathcal{O}(N)$, with the INDSCI-SIM model being slightly steeper due to the increased model complexity.

### S2 Appendix: Creating and benchmarking a synthetic population for Mumbai city

#### 2.1 Generating the Mumbai synthetic population

Located in the state of Maharashtra in India, Mumbai is one of India's largest cities, with a population of over 12 million. We use a subset of the IHDS-II dataset obtained by filtering for individuals and households which are situated in the state of Maharashtra. (We use the filtered datasets for Maharashtra because the Mumbai dataset in IHDS-II contains a few hundred samples, which are not enough to be able to generate a quality synthetic population.) Each job description is drawn from the empirical distribution observed in the IHDS-II subset for Maharashtra.

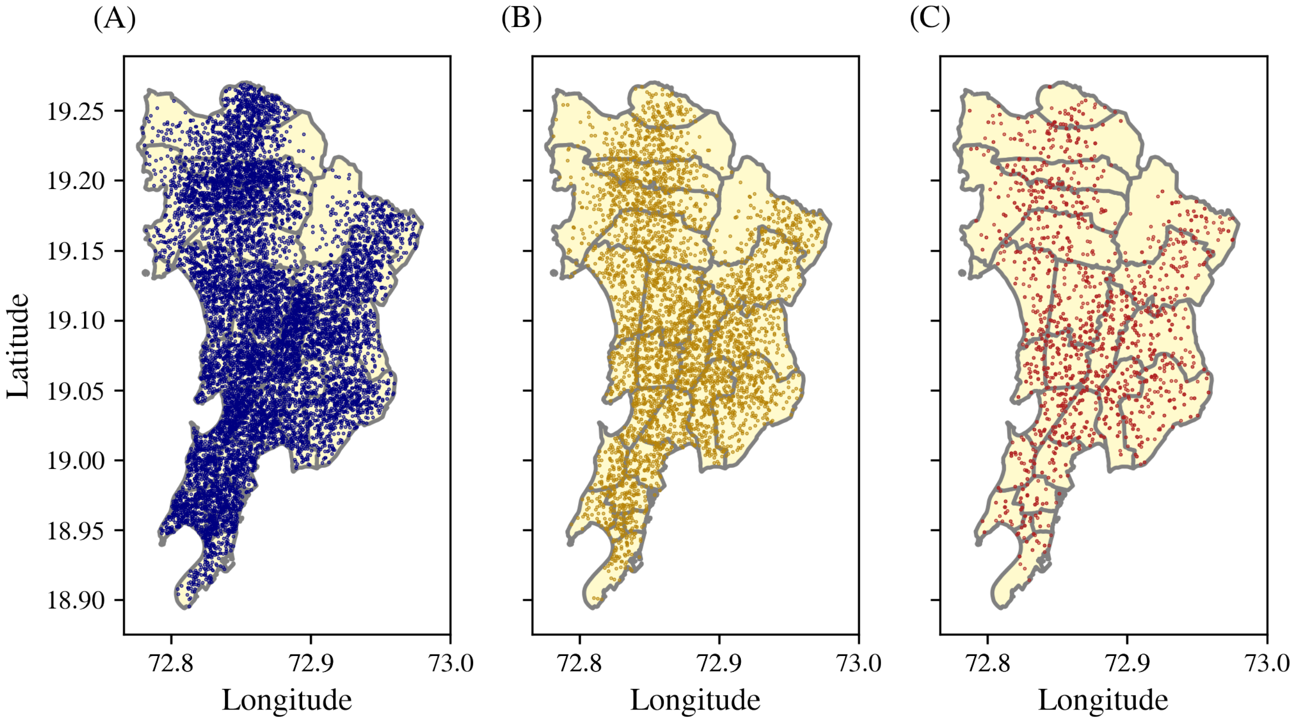


**Fig S2.1: Distribution of geo-locations.** Geographical distribution of (A) households, (B) workplaces, and (C) schools for the combined synthetic population of the districts of Mumbai and Mumbai Suburban. The underlying map of Mumbai is provided by the Spatial Data of Municipalities (Maps) Project by Data{Meet} [1].

Next, we generate synthetic workplaces, schools, and public places for our agents as detailed in Section 2.1.2 in the main paper. The final synthetic population has demographic data (age, height, and so on), disease data (chronic heart disease, diabetes, and other co-morbidities), family data, geographic location for workplaces, schools, and other geo-locations, and socio-economic data (job label, and religion). In total, we generate a synthetic population with 44 attributes. These attributes and their datatypes are shown in Table S2.1.

| Col | Title | Datatype | Col | Title | Datatype |
|---|---|---|---|---|---|
| 1 | AgentID | int64 | 23 | District | string |
| 2 | SexLabel | string | 24 | JobType | string |
| 3 | Age | int64 | 25 | EssentialWorker | bool |
| 4 | Height | float64 | 26 | AdminUnit_Name | string |
| 5 | Weight | float64 | 27 | AdminUnit_Lat | float64 |
| 6 | Religion | string | 28 | AdminUnit_Lon | float64 |
| 7 | Caste | string | 29 | HHID | int64 |
| 8 | M_Fever | bool | 30 | H_Lat | float64 |
| 9 | M_Cough | bool | 31 | H_Lon | float64 |
| 10 | M_Diarrhea | bool | 32 | AdherenceToIntervention | float64 |
| 11 | M_Cataract | bool | 33 | UsesPublicTransport | bool |
| 12 | M_TB | bool | 34 | WorkPlaceID | int64 |
| 13 | M_HighBP | bool | 35 | W_Lat | float64 |
| 14 | M_HeartDisease | bool | 36 | W_Lon | float64 |
| 15 | M_Diabetes | bool | 38 | WorkPlace_AdminUnit | string |
| 16 | M_Leprosy | bool | 38 | SchoolID | int64 |
| 17 | M_Cancer | bool | 39 | School_Lat | float64 |
| 18 | M_Asthma | bool | 40 | School_Lon | float64 |
| 19 | M_Polio | bool | 41 | School_AdminUnit | string |
| 20 | M_Paralysis | bool | 42 | PublicPlaceID | int64 |
| 21 | M_Epilepsy | bool | 43 | PublicPlace_Lat | float64 |
| 22 | StateLabel | string | 44 | PublicPlace_Lon | float64 |

**Table S2.1: Columns in the synthetic population.** The 44 attributes present in the created synthetic population and their datatypes are shown.

#### 2.2 Benchmarking our population against survey data

Fig S2.2 compares the marginal distributions of attributes of age, height, and weight across samples of the survey and the synthetic population for the combined districts of Mumbai and Mumbai Suburban. We work with a randomly chosen subset comprising 10,000 individuals to compare with the survey data, although our synthetic population has 12 million individuals.

Table S2.2 shows results for two statistical tests, the two-sample Kolmogorov–Smirnov (KS) test and the Chi-Squared (CS) test. These statistical tests are conducted on all the compatible columns in both the survey and synthetic population, so that the CS test is applied to categorical or Boolean columns, and the KS test is applied to numerical columns. In each case, we report a confidence level for each test that represents the confidence that the synthetic data and survey data come from the same distribution. For the CS test, the confidence level is the $p-$value, while for the KS test it is 1 - (KS statistic). The average confidence levels for relevant columns are reported in Table S2.2.

#### 2.3 Metrics for the synthetic population

To further study how faithfully our synthetic population represents the survey data from the IHDS-II dataset, we run a series of tests to compare their statistical features. The data in our population is of two types: numerical and categorical. We use different tests to check the similarity between

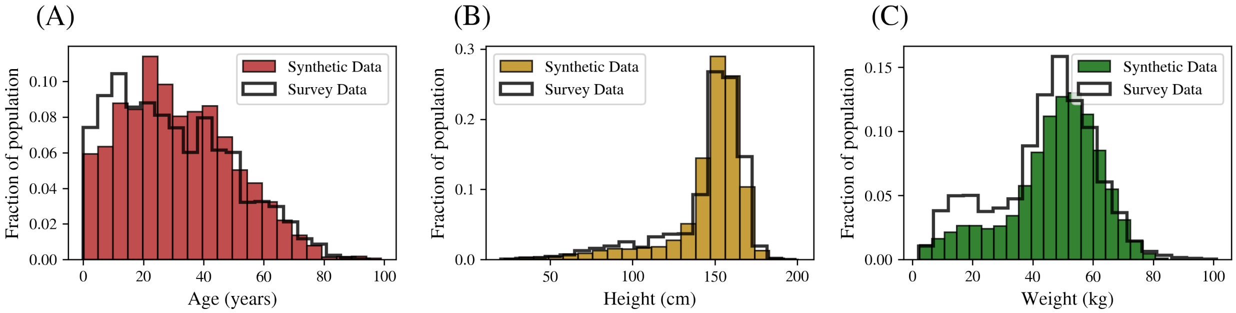


**Fig S2.2: Histograms of marginal distributions.** The distributions of age, height and weight in the synthetic population for the combined districts of Mumbai and Mumbai Suburban. In each case, this distribution is plotted over the distributions obtained from survey data, and these distributions compared using the Kolmogorov-Smirnov test. The results and KS statistic for each case are given in Table S2.2.

| **Test** | **Confidence level** |
|---|---|
| CS Test for Sex | 0.99 |
| KS Test for Age | 0.98 |
| KS Test for Height | 0.91 |
| KS Test for Weight | 0.95 |

**Table S2.2: Statistical benchmarks of the synthetic population.** To compare the distributions of various columns, we use the chi-square test for columns with discrete, categorical data (sex in our population). This test returns a $p-$value. A high $p-$value indicates the confidence that the synthetic data and the survey data come from the same discrete distribution. We also use the Kolmogorov-Smirnov statistic to compare the marginal distributions of continuous, numerical data (age, height, and weight in our population). We report a score of 1-(KS statistic). A higher score again indicates the confidence that the synthetic data and the survey data come from the same continuous distribution.

the generated population and the survey dataset. Three of these tests are for categorical columns: `CategoryCoverage`, `ContingencySimilarity`, and `TVComplement`, while the others are for the numerical columns. We report the results for the tests described below on columns that are present in both the generated synthetic population and the IHDS dataset. These results are shown in Tables S2.3 and S2.4. In every test a result of 1.0 signifies that the survey and synthetic population are identical, and 0.0 signifies that they survey and synthetic population are very dissimilar.

`CategoryCoverage` Checks whether the synthetic column covers all the categories present in the survey column.

`TVComplement` Computes the similarity between synthetic and survey categorical columns in terms of their marginal distributions.

`BoundaryAdherence` Checks whether a column from the synthetic data respects the range of the survey data or not. It does so by comparing the minimum and maximum values from both datasets.

`CorrelationSimilarity` Compares the trends of 2D distributions by measuring the Pearson correlation coefficient between a pair of numerical columns and computing the similarity between the survey and synthetic datasets.

`RangeCoverage` Checks whether the synthetic column covers the entire range of values present in the survey column.

`KSComplement` Computes the similarity between synthetic and survey numerical columns in terms of their marginal distributions.

`ContingencySimilarity` Compares the 2D distributions by computing the similarity of a pair of columns between the survey and synthetic categorical columns.

`StatisticSimilarity` Using mean, median and standard deviation it computes the similarity between a survey and synthetic numerical column.

| **Features** | BoundaryAdherence | RangeCoverage | StatisticSimilarity | KSComplement |
|---|---|---|---|---|
| Height | 0.9999 | 0.9879 | 0.9893 | 0.9474 |
| Weight | 1.0000 | 0.8400 | 0.9657 | 0.8907 |
| Age | 0.9990 | 1.0000 | 0.9851 | 0.9176 |

| **Feature** | TVComplement |
|---|---|
| SexLabel | 0.9296 |
| M_Cough | 0.9996 |
| M_Cancer | 0.9997 |
| M_Diarrhea | 0.9995 |
| M_Fever | 0.9997 |
| M_Cataract | 0.9992 |
| M_TB | 1.0000 |
| M_HeartDisease | 0.9987 |
| M_Diabetes | 0.9999 |
| M_HighBP | 0.9997 |
| M_Leprosy | 0.9998 |
| M_Asthma | 0.9996 |
| M_Paralysis | 0.9996 |
| M_Epilepsy | 0.9981 |
| M_Polio | 1.0000 |

**Table S2.3:** Metrics for comparing numerical (age, height, and weight) and categorical (comorbidity) columns between the Mumbai synthetic population and the IHDS-II survey data. In every test a result of 1.0 signifies strong correlation and 0.0 signifies no correlation between the survey and synthetic data.

We also consider joint tests for numerical columns to verify that correlations between the different columns are faithfully represented.

| Features | CorrelationSimilarity |
| --- | --- |
| Age, Height | 0.9999 |
| Age, Weight | 0.9838 |
| Height, Weight | 0.9754 |

| Features | ContingencySimilarity |
| --- | --- |
| M_Cough, M_HeartDisease | 0.9983 |
| M_Diabetes, M_HeartDisease | 0.9987 |
| M_Cough, M_Fever | 0.9994 |
| M_Cough, M_Asthma | 0.9994 |

**Table S2.4:** Metrics for comparing the joint distributions of numerical (age, height, and weight) and categorical (comorbidity) columns between the Mumbai synthetic population and survey data from the IHDS-II survey. In every test a result of 1.0 signifies strong correlation and 0.0 signifies no correlation between the survey and synthetic data.

### S3 Appendix: Description of the compartmental model

As mentioned in the main text, the INDSCI-SIM model contains 9-compartments. These are (S)usceptible, (E)xposed, Asymptomatic ($I^A$), Presymptomatic ($I^P$), Mildly Infected ($I^M$), Severely Infected ($I^S$), (H)ospitalized, (R)ecovered, and (D)ead. These are shown in Fig 5 of the main text. Furthermore, each of these compartments is age-stratified, as denoted by the subscript $i$. Using these definitions, the dynamics of our compartmental model can be represented by the following equations:

$$\begin{aligned}
\dot{S}_i &= -\beta S_i \sum_{j=1}^{N_{\text{age}}} I_j^{\text{eff}} + \zeta R_i \\
\dot{E}_i &= \beta S_i \sum_{j=1}^{N_{\text{age}}} I_j^{\text{eff}} - \gamma E_i \\
\dot{I}_i^A &= \alpha \gamma E_i - \lambda_A I_i^A \\
\dot{I}_i^P &= (1-\alpha)\gamma E_i - \lambda_P I_i^P \\
\dot{I}_i^M &= \mu \lambda_P I_i^P - \lambda_M I_i^M \\
\dot{I}_i^S &= (1-\mu)\lambda_P I_i^P - \lambda_S I_i^S \\
\dot{H}_i &= \lambda_S I_i^S - \rho H_i \\
\dot{R}_i &= (1-\delta)\rho H_i + \lambda_A I_i^A + \lambda_M I_i^M - \zeta R_i \\
\dot{D}_i &= \delta \rho H_i
\end{aligned} \tag{1}$$

where the parameters are described in Table S3.1, and $I_j^{\text{eff}}$ is given by:

$$I_j^{\text{eff}} = \left(C_A I_j^A + C_P I_j^P + C_M I_j^M + C_S I_j^S\right)/N_j.$$

| Parameter | Description |
|---|---|
| $\beta$ | Rate at which infected individuals can infect the susceptible population |
| $\gamma$ | Rate at which exposed individuals become infectious |
| $\lambda_A$ | Asymptomatic individuals remain infected for an average of $1/\lambda_A$ days |
| $\lambda_P$ | Presymptomatic individuals remain infected for an average of $1/\lambda_P$ days |
| $\lambda_M$ | Individuals experiencing mild symptoms recover on an average in $1/\lambda_M$ days |
| $\lambda_S$ | Individuals experiencing severe symptoms are hospitalized on an average in $1/\lambda_S$ days |
| $\rho$ | Hospitalized individuals remain hospitalized for an average of $1/\rho$ days |
| $\zeta$ | Recovered individuals remain immune to the disease for an average of $1/\zeta$ days |
| $\alpha$ | Fraction of exposed individuals who are asymptomatic carriers of the disease |
| $\mu$ | Fraction of presymptomatic carriers who become mildly infected |
| $\delta$ | Fraction of hospitalized individuals who die from the disease |
| $C_A$ | Contact parameter: relative risk of an asymptomatic agent infecting a susceptible agent |
| $C_P$ | Contact parameter: relative risk of a presymptomatic agent infecting a susceptible agent |
| $C_M$ | Contact parameter: relative risk of a mildly-infected agent infecting a susceptible agent |
| $C_S$ | Contact parameter: relative risk of a severely-infected agent infecting a susceptible agent |

**Table S3.1:** Description of the parameters used in Eq S1.

However, it is important to stress that we only use the compartments described in this model to label the different states in the disease progression. The dynamics described in the paper differ significantly from those of the compartmental model above in two significant ways: (i) our model in the paper has multiple network interactions and therefore incorporates differential contacts between agents, both spatially and temporally, and (ii) the sojourn times of agents in the different disease compartments of our model are taken to be lognormally distributed (see Table S3 of the main paper), while those of the well-mixed compartmental model are exponentially distributed.

#### 3.1 On the equivalence of compartmental and well-mixed agent-based model

In this supplementary section, we show that we can closely reproduce results from the above compartmental model using our agent-based simulation. We do this by (i) simulating a well-mixed scenario in which every agent interacts with with each other agent at any given time, and (ii) incorporating exponential, rather than lognormally distributed, residence times.

In Fig S3.1 we show the results for the total number of active infections in an agent-based simulation (averaged over multiple stochastic runs), and compare it to the results obtained by solving the differential equations in Eq S1. These simulations are run using the parameters defined in Table S3.2.

| Parameter | Value | Parameter | Value | Parameter | Value |
|---|---|---|---|---|---|
| $C_A$ | 1.0 | $\gamma$ | 1/5 | $\rho$ | 1/18 |
| $C_P$ | 1.0 | $\lambda_A$ | 1/5 | $\zeta$ | 0.0 |
| $C_M$ | 1.0 | $\lambda_P$ | 1/2 | $\alpha$ | 0.35 |
| $C_S$ | 1.0 | $\lambda_M$ | 1/8 | $\mu$ | 0.9792 |
| $\beta$ | 0.3 | $\lambda_S$ | 1/2 | $\delta$ | 0.00032 |

**Table S3.2:** Parameter values used in comparing the agent-based and differential equation model (described by Eq S1).

In Fig S3.1 we show the epidemic curves for all infected individuals in the population, both using our agent-based simulator and using the differential equation based compartmental model described in Eq S1. We see from the figure that while a small difference exists between the results from the compartmental model and those of our agent-based simulations, this difference reduces as the time-step of the agent-based simulation is reduced. This difference, that we attribute to the discrete-time nature of our agent-based simulator, can be accounted for by a small change in the parameter $\beta$.

In Fig S3.2, we show the results for all compartments in the disease progression, from which we conclude that our agent-based simulation does indeed agree with the results from the compartmental model. We show further in Fig S3.3 that a perfect matching between the agent-based and compartmental results can be obtained by allowing a single parameter ($\beta$) to vary by less than 5% (i.e. changing $\beta = 0.3$ to $\beta = 0.31$).

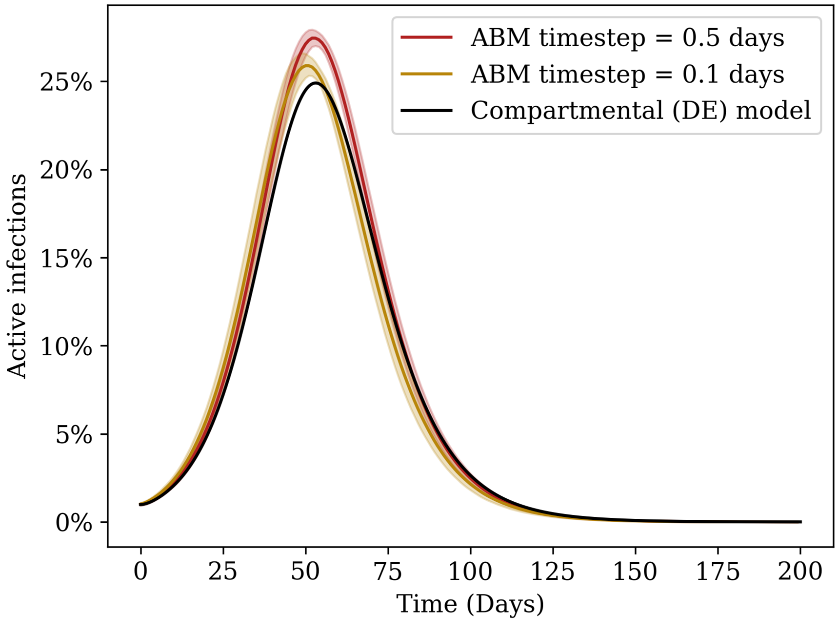


**Fig S3.1: Comparing agent-based and differential-equation based models.** Epidemic curves showing the total infected individuals in the population as a function of time. The red curve shows the results using a time-step of half-a-day in our agent-based simulations, while the golden curve shows the results using a time-step of one-tenth of a day. In each case, a population of 100,000 well-mixed individuals was considered, and 200 simulations were run and averaged over. The shaded regions represent error bars of $1.96\sigma$. The black solid curve shows the same curve for the compartmental model, obtained by solving the differential equations in Eq S1 with the same initial conditions. While the results of our model differ marginally from those of the compartmental model, we attribute this to the discrete time-step used. Indeed, as the time-step is lowered, a stronger agreement is seen. The models can be made to match exactly if the parameter $\beta$ is slightly changed, to account for the discrete-time effects.

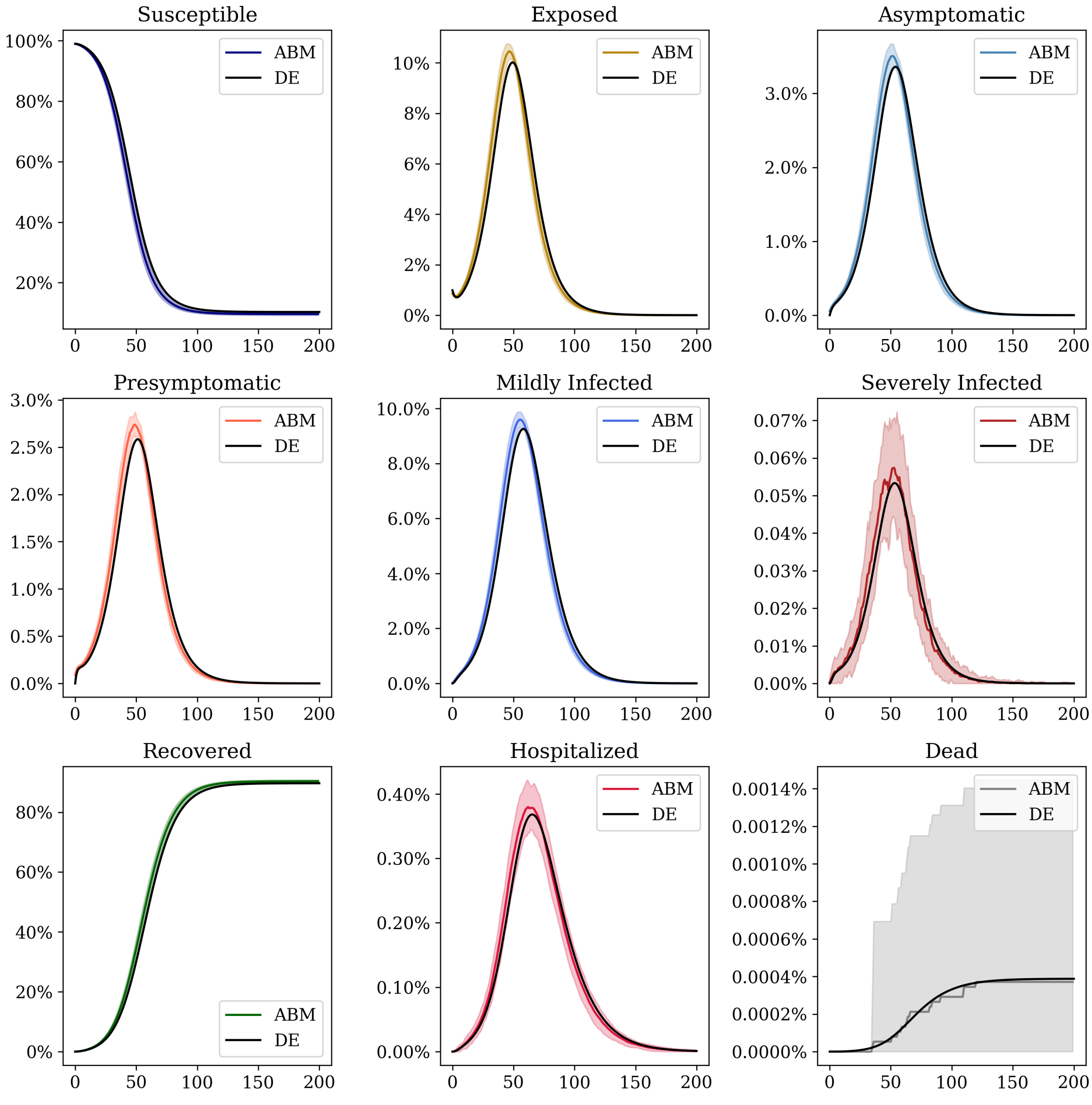


**Fig S3.2: Comparing agent-based and differential-equation based models.** Epidemic curves for each compartment showing the total infected individuals in the population as a function of time. In each case, a population of 100,000 well-mixed individuals was considered, and 200 simulations were run and averaged over. The shaded regions represent error bars of $1.96\sigma$. The black solid curve shows the same curve for the compartmental model, obtained by solving the differential equations in Eq S1 with the same initial conditions.

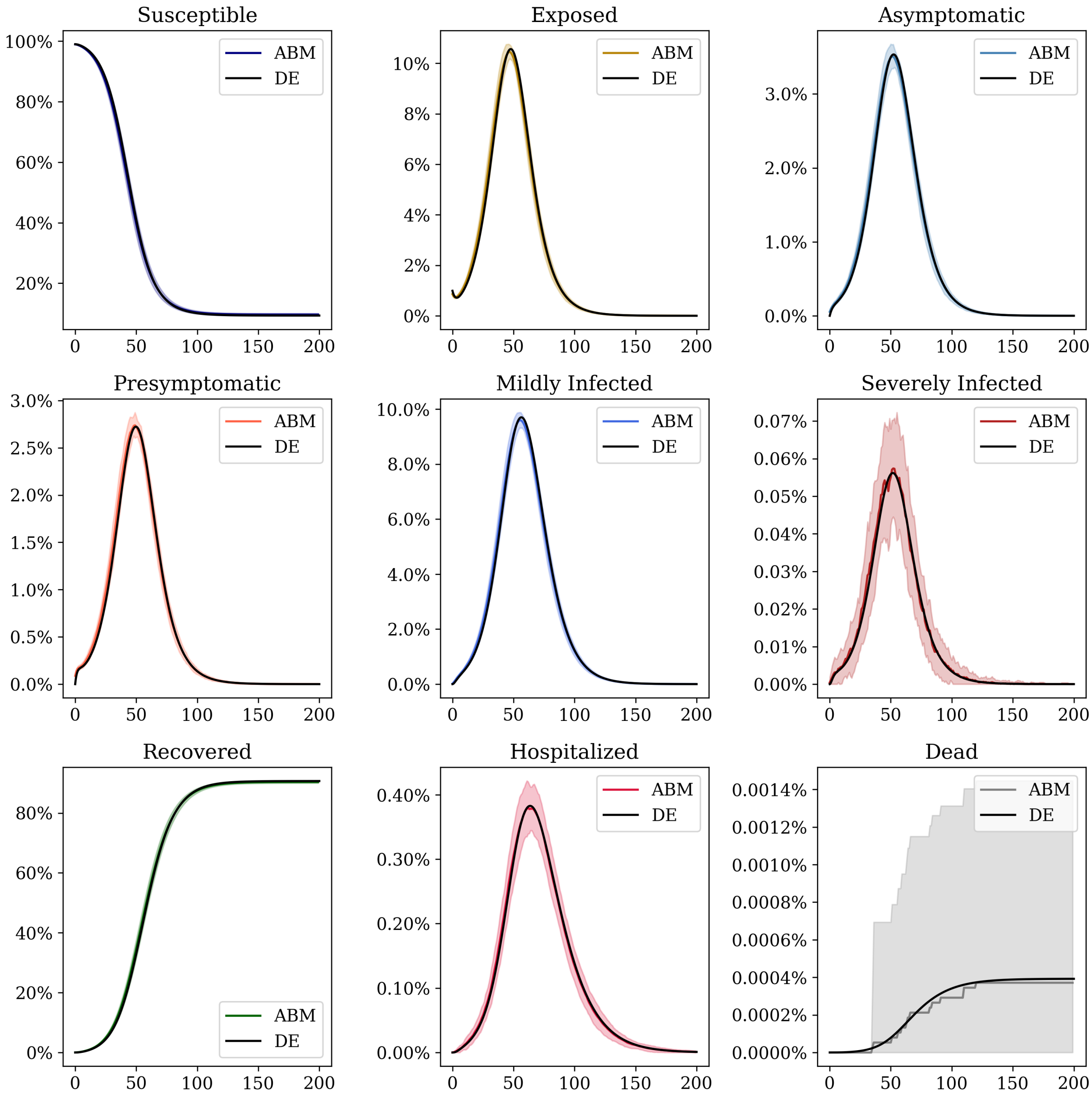


**Fig S3.3: Accounting for discrete-time effects.** The same graphs as in Fig S3.2, but using a value of $\beta = 0.31$ in the compartmental model. We can see that this small change in $\beta$ makes all curves from our ABM simulation agree with their compartmental counterparts.

#### S4 Appendix: Sensitivity analyses

Here we discuss the sensitivity of the results discussed in the main paper to some of our model choices.

##### 4.1 Sensitivity to home-workplace travel distance

In order to study the role played by the geographical structure of our network, we construct multiple populations for the city of Pune in which individual agents are preferentially assigned workplaces closer to their homes. In Fig S4.1A we show the distribution of the "travel distances" for the agents in our population.

We run simulations with these populations, and for a range of values of the parameter $\beta$, which modulates the transmissibility of the disease. In each case we compute the "outbreak size", i.e. the total number of individuals who contracted the disease over the duration of the epidemic. This number is averaged over multiple stochastic runs and the result is plotted in Fig S4.1B. As can be seen from the graphs, the distribution of travel times has a very low effect on the outbreak size. We have also repeated this process for the epidemic curves. We have verified that the difference between them in all the scenarios discussed in the main paper is marginal.

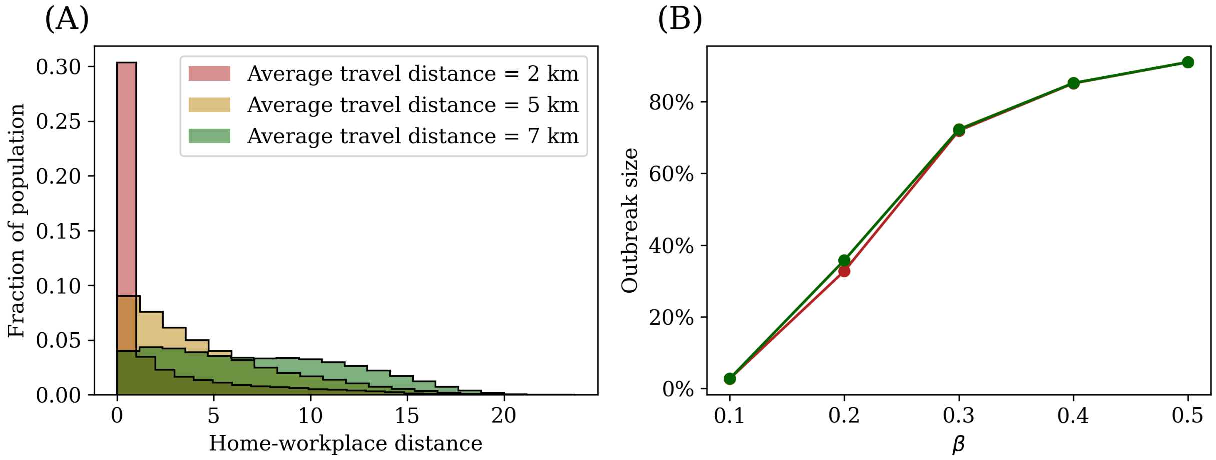


**Fig S4.1: Varying individual agents' travel distances.** We run our simulations on three different populations in which which we vary the distance that individuals travel between their home and work locations. In (A) we show the distribution of these travel-distances for all agents in each population. Populations in which this distribution is peaked at lower distances are those in which individuals are more likely to interact with other agents who are geographically close to their own homes. In (B) we show the outbreak size as a function of the tranmissibility for reach of these populations. As can be seen, as the transmissibility increases, so does the outbreak size. However, we note no significant difference in the results using the different populations. Each data point is the average over 10 simulation runs. Error bars are present at $1.96\sigma$, but are too small to be visible for this population size.

#### 4.2 Sensitivity to workplace occupancy

We further study the role that the distribution of workplace sizes on our results. In order to do this, we consider a population of 100,000 agents with varying average workplace occupancies. We vary both the workplace occupancy and the transmissivity of the disease and compute the outbreak size, as before, and average over multiple stochastic runs. Our results are shown in Fig S4.2. We see that beyond a certain value, the outbreak size is only weakly dependent of the workplace occupancy. Indeed, our results would hold for even workplaces that are as small as 50 agents. We have seen similar results in the literature; compare, for example Fig S4.2B with Fig. 4a(iii) in Ref [2].

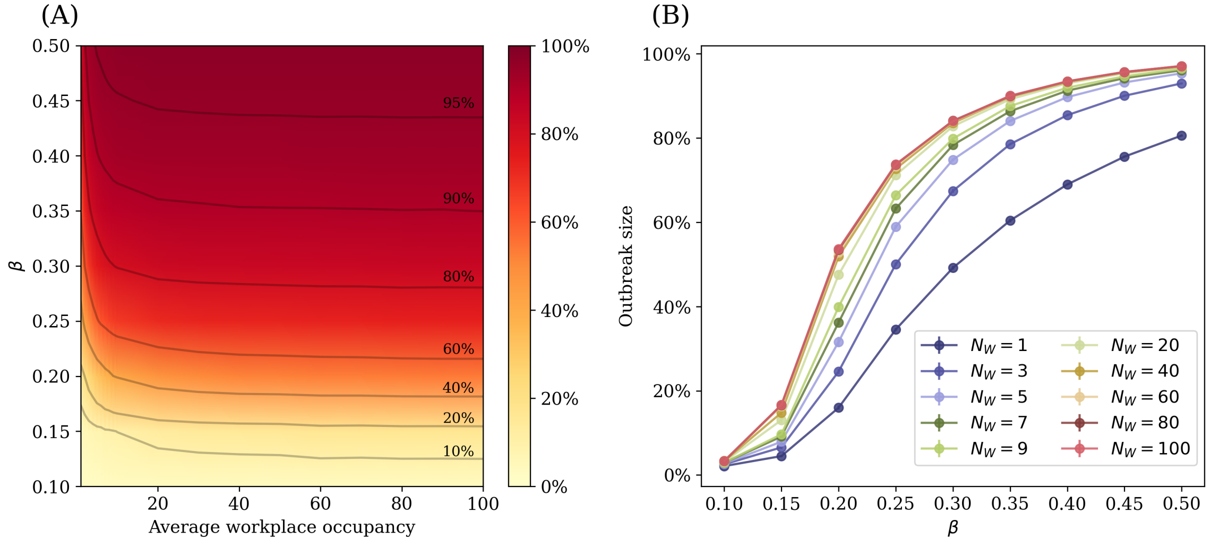


**Fig S4.2: Role of workplace occupancy in disease transmission.** We study the effect that workplace occupancy has on the transmission of the disease in our models. In (A) we plot a heatmap of the outbreak size as a function of the transmissibility $\beta$ and the workplace occupancy. Contours are plotted at different outbreak sizes. We see that the outbreak size is much more sensitive to $\beta$, and that beyond a workplace occupancy of around 50, the sensitivity to workplace size is very low. In (B) we show a subset of the same results, with each curve representing a single workplace occupancy. Each data point is the average over 200 simulation runs. Error bars are present at $1.96\sigma$, but are too small to be visible for this population size.

#### 4.3 Sensitivity to time spent at home

In order to quantify the sensitivity of our simulations to the time that agents spend at home, we run multiple simulations on a population of 100,000 agents, varying the number of time-steps spent at home. In order to do this, we run simulations in which the time-steps are 6 hours each (four time-steps in a day) and vary the number of steps that individual agents spend at their homes. In Fig S4.3 we show the results for the outbreak size of the simulations. We see that as the amount of time per day spent at home is increased, the outbreak size is reduced, although this is less significant both at low values of tranmissivity $\beta$ (when the infection dies out relatively quickly) and for high values of $\beta$ (when the infectious spreads very rapidly though the population), as is intuitively expected.

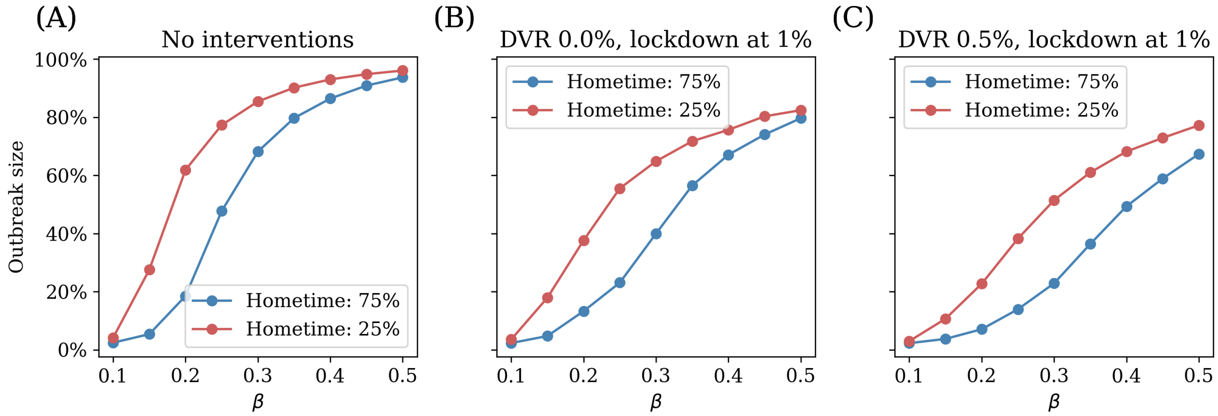


**Fig S4.3: Role of time spent at home and work locations on disease spread.** We investigate how our results would change based on the repartition of time spent in the low density (home) and high density (workplace) locations. We run multiple simulations with 6-hour time-steps, and vary the number of time-steps spent at home. We show the results here for 1 time-step spent at home (25% of the day, the red curves) and 3 time-steps spent at home (75% of the day, the blue curves). In each case, we compute the outbreak size, and plot it as a function of the transmissibility $\beta$. In (A) we show the results for the case where no interventions are applied to the population. In (B) and (C) we show results for when a lockdown is imposed when the number of active cases is 1% of the total population and with no vaccination drive, and with a daily vaccination rate of 0.5%. We see that the introduction of interventions like a lockdown and vaccination drive cause an overall reduction in the outbreak size, as we would expect. Each data point is the average over 50 simulation runs. Error bars are present at $1.96\sigma$, but are too small to be visible for this population size.

### S5 Appendix: The effect of vaccination

We consider vaccination as having three distinct effects: a reduction in the relative risk of infection, a reduction in the probability of severe infection, and a lower probability of transmitting the disease.

Vaccinated individuals are assumed to be 40% less likely to transmit the disease from the day they receive their first dose. The other relative effects evolve as a function of time. In both cases, we assume that the maximum protection from each dose is attained within 14 days of receiving the dose. In the interim period, we assume each parameter varies linearly. Fig S5.1A shows the reduction in the relative risk of infection, and Fig S5.1B shows the increase in the probability that a vaccinated individual will not exhibit symptoms.

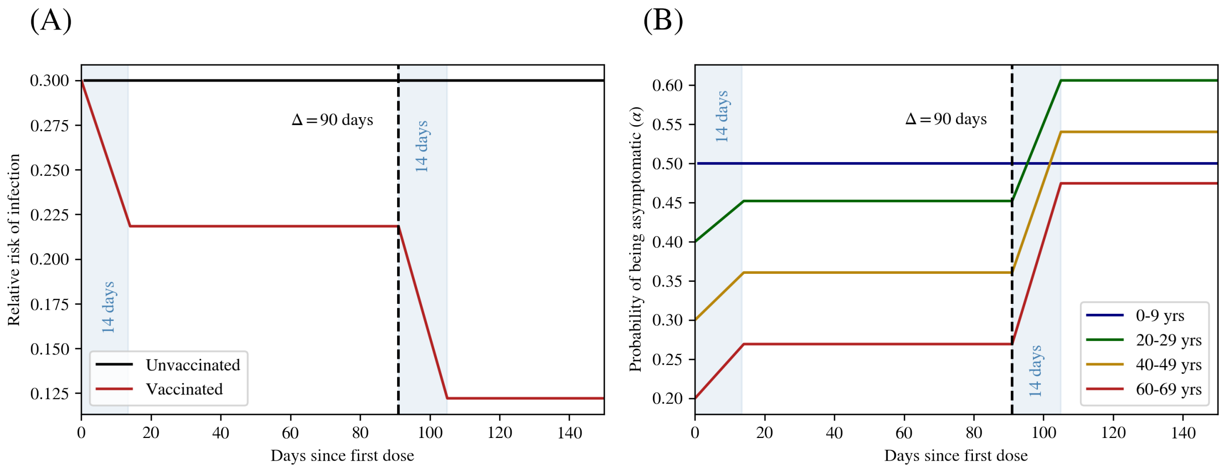


**Fig S5.1: Variation of relative risks of infection and severe disease as a function of time.** After each vaccine dose, these parameters reduce over a period of 14 days until the maximum protection from that dose is obtained. The same process is repeated after the second dose is obtained. (A) Reduction in the relative risk of infection reduces over a period of 14 days. (B) Increase in the probability of being asymptomatic.

#### S6 Appendix: Vaccine dose prioritization and avoiding wastage

The number of available vaccine doses that can distributed each day is decided by the daily vaccination rate. The allocation of vaccine doses is done by assuming some prior dose prioritization: some fraction $\tau$ of the total doses (chosen in our simulations to be 80%) are designated as "first" doses, and distributed amongst those individuals who are eligible to receive their first vaccine dose. The remaining fraction $(1-\tau)$ of doses are used to vaccinate individuals who are eligible for the second vaccine dose. However, this fraction is adjusted dynamically such that dose wastage is minimized.

At every time-step, a fixed number of individuals (set by the total number of available vaccines) is chosen from those who are eligible for the first shot of the vaccine. Individuals are eligible for vaccination provided (i) their age-band is one of those being vaccinated during the current phase of the vaccination drive, (ii) they have not yet received this specific dose, and (iii) the individual is not exhibiting symptoms. The same process is repeated for the second dose. For the second dose of the vaccine, in addition to the earlier eligibility criteria, at least $\Delta$ days must have elapsed since the individuals received their first dose. In our simulations we choose $\Delta = 90$.

Once these individuals are chosen, fractions $\tau$ and $(1-\tau)$ of the vaccines are administered to those eligible to receive the first and second doses respectively. If there are any leftover vaccines (for example, there are more first doses available that eligible individuals), we then evaluate the excess doses for both first and second shots, swap them, and re-administer. In other words, we use any excess second doses to vaccinate those eligible for their first dose and vice versa. While the resulting ratio will not necessarily be $\tau$, this ensures that all available doses are used in a single day, unless there are no eligible people in the population for any vaccine dose.

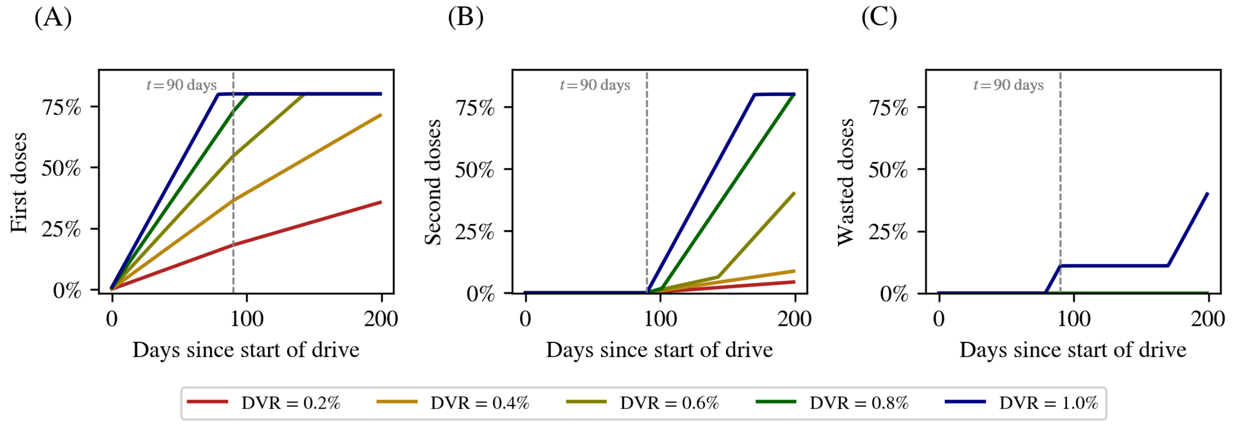


**Fig S6.1: First, second, and wasted doses as a function of time.** The fraction of the total population that are vaccinated with (A) first and (B) second doses for daily vaccination rates from 0.2% to 1%. In all cases, the vaccine drive starts on day 0. The dose-prioritization ratio is 80:20, with 20% of the doses being designated as second-doses. For the first 90 days, all doses administered are first-doses, since no one is eligible for a second dose, after which second doses start getting administered. The numbers plateau at a value less than 100% since not everyone in the population is eligible for a vaccine. In (C) we show the number of wasted doses, again as a fraction of the total population. In the case of DVR = 1%, the vaccination rate is so high that there are occasionally more doses available than eligible agents, leading to dose wastage.

### S7 Appendix: Descriptions of the synthetic populations used in our simulations

We describe the synthetic population used in our simulations for the city of Pune, and for the study of school reopenings.

#### 7.1 Metrics for the synthetic population of Pune

In Fig S7.1 we show the geographical distribution of homes, workplaces, and schools.

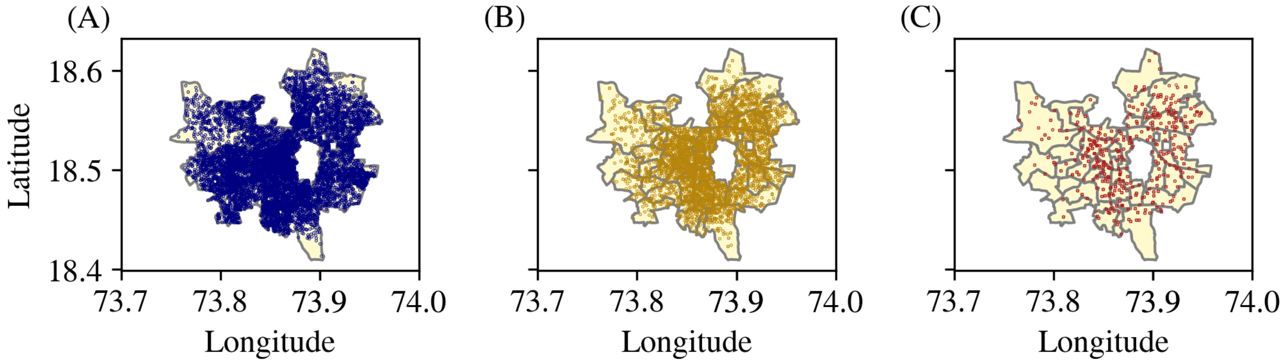


**Fig S7.1: Distribution of geo-locations for the city of Pune.** Geographical distribution of (A) households, (B) workplaces, and (C) schools for the combined synthetic population of the districts of Pune. The underlying map of Pune is provided by the Spatial Data of Municipalities (Maps) Project by Data{Meet} [1].

In Fig S7.2, we compare the distribution of ages in the survey and the synthetic population of Pune. We work with a randomly chosen subset of the synthetic population comprising 10,000 individuals to compare with the survey data, although our full synthetic population has over 3 million individuals.

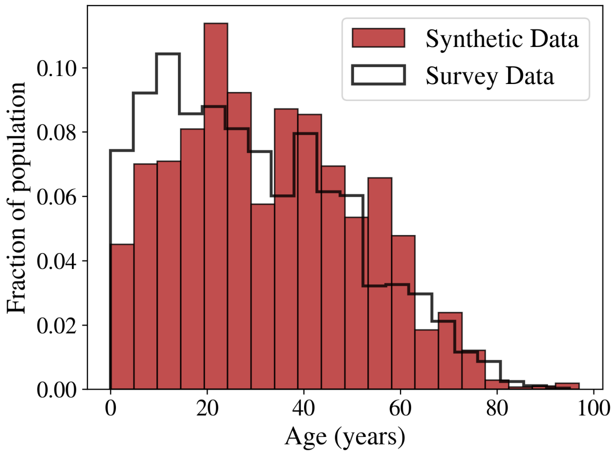


**Fig S7.2: Histogram of age distribution.** The distributions of age in the synthetic population for the population of Pune for both the synthetic population used in the modelling results of the main paper, and data from the IHDS-II survey.

We also compute statistical metrics, as described in Appendix S2, to compare our population with the IHDS-II dataset for Maharashtra. The results are shown in Tables S7.1 and S7.2.

| **Features** | Boundary Coverage | RangeCoverage | StatisticSimilarity | KSComplement |
|---|---|---|---|---|
| Height | 1.0000 | 0.9000 | 0.9800 | 0.9000 |
| Weight | 1.0000 | 0.6300 | 0.9600 | 0.7400 |
| Age | 1.0000 | 1.0000 | 0.9700 | 0.8900 |

| **Feature** | **TVComplement** |
|---|---|
| SexLabel | 0.9200 |
| M_Cough | 0.9980 |
| M_Cancer | 0.9990 |
| M_Diarrhea | 0.9990 |
| M_Fever | 0.9990 |
| M_Cataract | 0.9980 |
| M_TB | 1.0000 |
| M_HeartDisease | 0.9990 |
| M_Diabetes | 0.9990 |
| M_HighBP | 0.9990 |
| M_Leprosy | 0.9990 |
| M_Asthma | 0.9990 |
| M_Paralysis | 0.9990 |
| M_Epilepsy | 0.9980 |
| M_Polio | 1.0000 |

**Table S7.1:** Metrics for comparing numerical (age, height, and weight) and categorical (comorbidity) columns between the Pune synthetic population used in our simulations and the IHDS-II survey data. In every test a result of 1.0 signifies strong correlation and 0.0 signifies no correlation between the survey and synthetic data.

#### 7.2 Description of the school population

A section of the synthetic population for the city of Pune is chosen with 20,316 individuals with 6500 homes, 120 workplaces, and 1 school. These individuals represent the catchment area for the school. In Fig S7.3 we show the geographical distribution of the school population, and in Fig S7.4 we show some of its statistics.

| Features | CorrelationSimilarity |
|---|---|
| Age, Height | 0.9900 |
| Age, Weight | 0.9800 |
| Height, Weight | 0.9500 |

| Features | ContingencySimilarity |
|---|---|
| M_Cough, M_HeartDisease | 0.9980 |
| M_Diabetes, M_HeartDisease | 0.9990 |
| M_Cough, M_Fever | 0.9980 |
| M_Cough, M_Asthma | 0.9980 |

**Table S7.2:** Metrics for comparing the joint distributions of numerical (age, height, and weight) and categorical (comorbidity) columns between the Pune synthetic population used in our simulations and survey data from IHDS-II. In every test a result of 1.0 signifies strong correlation, and 0.0 signifies no correlation, between the survey and synthetic data.

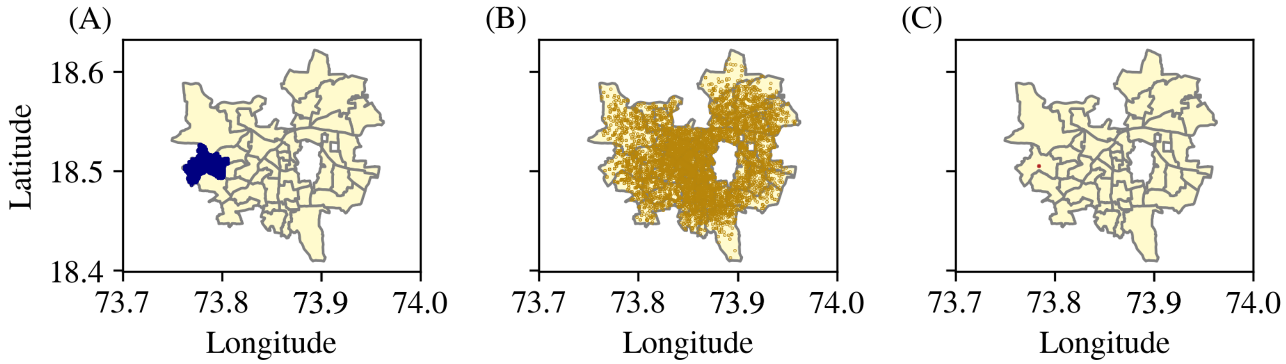


**Fig S7.3: School synthetic population geographical distribution.** The distributions of (A) households, (B) workplaces, and (C) (one single) school for this section of the population. The underlying map of Pune is provided by the Spatial Data of Municipalities (Maps) Project by Data{Meet} [1].

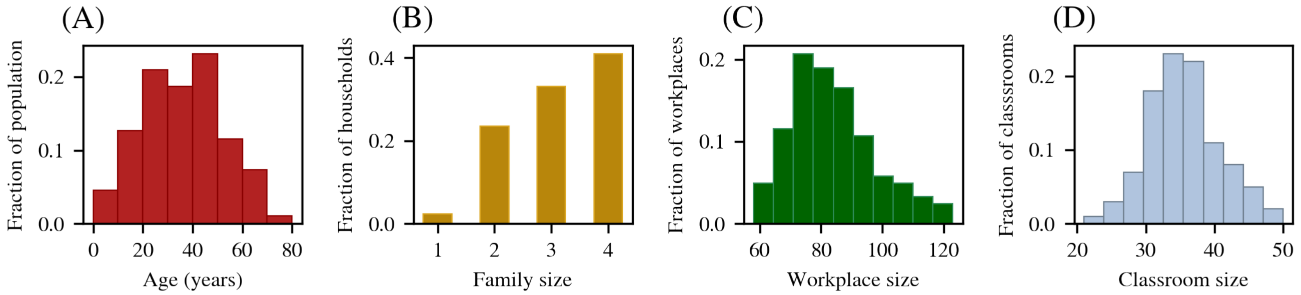


**Fig S7.4: School synthetic population statistics.** The distributions of (A) age, (B) family size, (C) workplace size, and (D) classroom size for this section of the population.

### S8 Appendix: Using BharatSim for epidemic forecasts: a case study for Pune

In this section, we describe how BharatSim can be used for epidemic forecasts. We use real data for confirmed cases from the initial part of the first wave of COVID-19 in the Indian city of Pune. In the period we consider, the city was twice placed under a rigorous lockdown, with an intervening period of several weeks when the lockdown was lifted.

Here we demonstrate the versatility of BharatSim by comparing model projections based on initial independent estimates of the basic reproductive ratio $R_0$, to real data. The collective network properties of agent interactions are modified by public health interventions such as lockdowns. The projections, and an understanding of how they are modulated by the interventions made, can then be directly used in public health planning in an epidemic situation.

We first calibrate the value of $\beta$ used in our simulations to match the initial $R_0$. We obtain $R_0$ from a statistical analysis of daily case data [3]. These estimates can be further refined as time proceeds.

#### 8.1 Computing the reproductive ratio from our simulations

The basic reproductive ratio is defined as the average number of people infected by a single contagious person in the background of a population of susceptible people. Its value, in an averaged sense, depends on the mean number of contacts, the probability that an infection will result from any given contact over the period of the interaction, and the time over which an individual remains infected. In our agent-based simulations, network structure plays an important role in determining $R_0$.

To compute this quantity from our simulation results, we keep a record of which agent was responsible for each infection. Whenever an individual is infected in a specific location, an agent is drawn at random from the potential "infectors" at that location so that infection spread can be described.

The simulations are run for a range of values of $\beta$, for a period of 21 days. This period is chosen so that a sufficient number of infectious individuals recover. At the end of this period, the average number of people infected by a person who has recovered from the disease, is computed. These results are shown in Fig S8.1. We conclude from this figure that if we require a value of $R_0$ to be around 1.2–1.3, as estimated in the statistical analysis of case data for this period (see Section 6.4 in the Supplementary Information of Ref [4]), a value of $\beta = 0.4$ should be used in our simulation.

#### 8.2 Predicting the spread of COVID-19 in the first wave

We begin our simulations with our population in a lockdown state. The initial lockdown in Pune was imposed from the 25th of March until the 31st of May, 2020. During this period, the agents in our simulation were allowed to have only limited mobility – only those marked as "essential workers" were allowed to move out of their homes. We began the simulation by seeding 1500 exposed agents in the population, distributed randomly around the city. After the 31st of May, the lockdown was lifted, and agents allowed to move between their homes and workplaces. This led to an increase in the spread of the disease. On the 14th of July, a second ten-day lockdown was imposed in Pune.

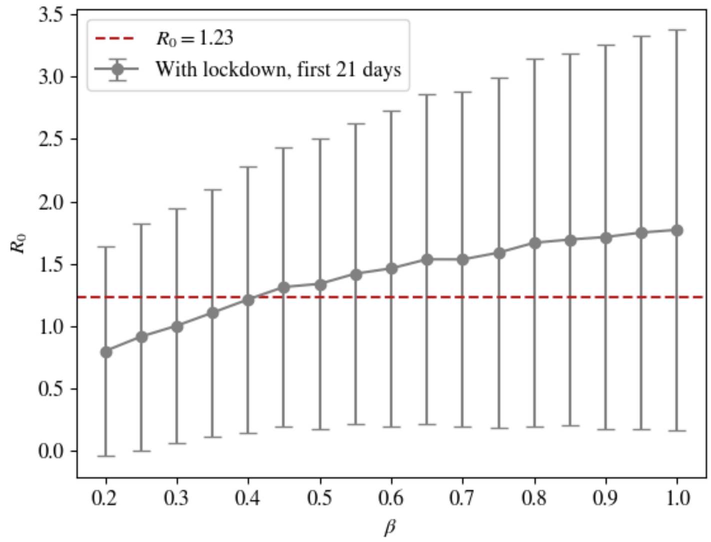


**Fig S8.1:** The basic reproductive ratio obtained from our simulation as a function of the transmission factor $\beta$. We see that in order to obtain an $R_0$ in the range 1.2 to 1.3, a value of $\beta = 0.4$ must be used.

During the entire simulation, schools remained closed, as was the situation across the first wave. Our simulations thus capture the actual interventions that were applied, and can be used to model the response in the numbers of infected to those interventions.

##### 8.3 Results

In Fig S8.2 we show our results by comparing the number of daily recorded cases with the number of active symptomatic cases in the population, corrected for under-counting. (Due to major limitations in testing during much of the first wave of COVID-19 in India, as well as social stigma associated with reporting infections, it is believed that case under-counting was substantial in the first several months of the spread of COVID-19 in India.) We account for case under-counting by dividing the total number of active symptomatic cases estimated in the simulation by a constant factor of 150; this is consistent with national estimates for India in the same period.

Fig S8.2 shows that, with our initial conditions, we are able to predict the spread of the disease quite accurately until the end of the second lockdown, given only (a) an estimate for initial infections, (b) a single number representing the over-counting factor and (c) a value of $R_0$ extracted from a time-series analysis of the first few weeks of reported cases. It is difficult to project beyond this, since the mix of variants changed decisively across the month of August 2020 [5,6], and reinfections arising from waning immunity from a prior infection (which we excluded in this analysis) became important. Nevertheless, our methods capture the rise in cases after the lockdown was lifted, the peak in cases during the second lockdown and the subsequent decrease in case numbers, attesting to the value of well-bench-marked agent-based models in providing real-time information regarding the trajectory of infections during an ongoing epidemic.

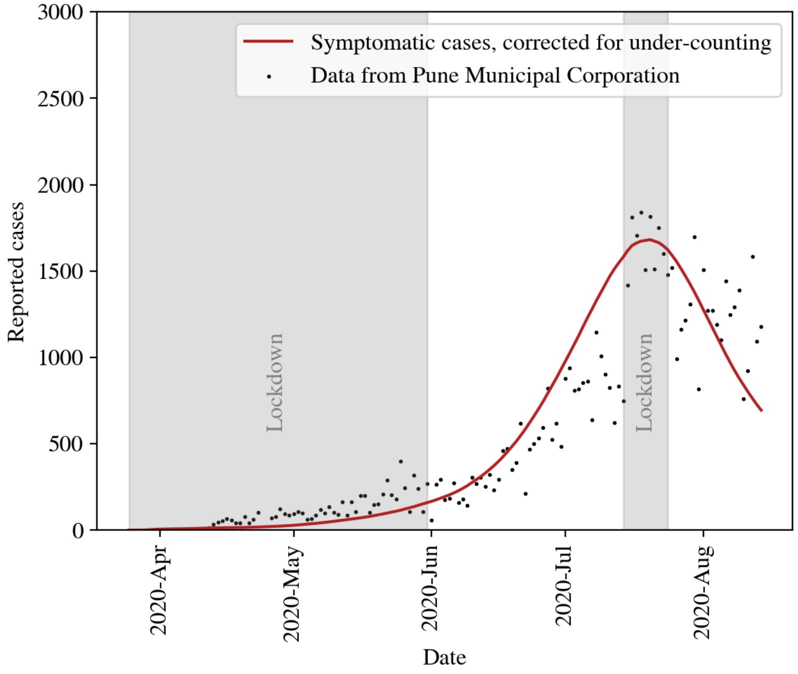


**Fig S8.2:** Our simulation results compared with data from the Pune Municipal Corporation. Using the value of $\beta = 0.4$ and an initial number of 1500 exposed agents, we are able to predict the spread of the disease, assuming a constant under-counting factor of 150. Our curves are obtained by looking only at the number of individuals in the symptomatic compartments of our model. If all infected individuals had been used, a similar result could be obtained, albeit with a higher under-counting factor.

### S9 Appendix: Studying the spread of mpox

Mpox is a viral infection caused by the mpox virus, a zoonotic virus in the genus *Orthopoxvirus*. Concerned by the sudden global spread of mpox beyond those African countries where it had previously been reported, the WHO declared it a Public Health Emergency of International Concern (PHEIC) in July, 2022. In the period between 01 January, 2022, and August 31, 2024, 123 countries have reported a total of 1,06,310 laboratory confirmed mpox cases and 234 deaths [7]. In India, 30 mpox cases have been reported since the WHO's 2022 PHEIC declaration [8].

Prior to the current outbreak, mpox infections were generally thought to arise mainly from contact with animal reservoirs. However, human-to-human transmission through direct routes (skin-to-skin contact, bodily fluids, and respiratory droplets) have also been recorded. Transmission could plausibly occur through sexually associated exposure to skin lesions, droplets, and fomites [9]. The 2022 outbreak of mpox was primarily associated with close intimate contact (including sexual activity) and most cases were diagnosed among men who have sex with men (MSMs), with 98% of the patients in a report of 528 cases from 16 countries being MSMs [10, 11].

In this section, we use BharatSim to model the spread of mpox in a community of MSMs and their associated household and workplace contacts.

#### 9.1 Creating a synthetic population

Since granular data for sexual contacts for MSMs is hard to obtain (since it might place individuals belonging to a stigmatised population at risk), our model bases itself on reconstructing sexual networks from aggregate data made available in [12] and [13].

As in the case of our simulations for multiple strains in the main paper (Section 3.3), we consider a population of 10,000 individuals, which we take to represent both MSMs and the network of their contacts. In principle, this could have been done with the BharatSim synthetic population directly, but the simpler procedure described here suffices to extract features of mpox spread in model populations with contact patterns similar to those characterised in the Indian context, without the overhead of the many additional (and for these purposes, irrelevant) variables that enter the description of the BharatSim population.

Of these 10,000 individuals, 1% are assumed to be MSM, while the others are their household and workplace contacts. The households and workplaces are distributed so that their mean occupancies are 4 and 50 respectively. This is done by first creating 2500 homes (corresponding to an average household size of 4 individuals) and assigning each individual to one of them with a uniform probability. The same process is repeated with workplaces, ensuring the correct average number of agents per location, distributed as a Poisson distribution.

#### 9.2 Assigning a contact network for MSMs

A quantity of some importance is the network of MSMs in the population. Studies have shown [9] that the sexual-networks of MSMs are often heavy-tailed, and are reasonably well modelled by distributions like the Weibull distribution. We use the range of number of partners shown in Ref [12], modeling this through a Weibull distribution, where the lowest and highest values in this range correspond to the 10% and 90% percentile respectively. The MSM network is created with this degree distribution in the following way:

- First, a trial number of contacts is chosen from the list of MSM agents. These numbers will be the starting point for our final distribution of contacts.
- Next, a list of all agents is made, with each agent being replicated as many times as they have contacts. This list is then shuffled to avoid the same agent being placed next to itself.
- Last, successive pairs of agents in the newly shuffled list are paired together, as far as possible, while avoiding mapping an agent as their own contact.

This method provides networks whose statistical properties are consistent with those available in the literature for MSM interactions.

The resulting distribution of contacts for MSMs and their graph is shown in Fig S9.1. This should be compared to similar figures in Ref [12].

#### 9.3 The disease progression

Mpox is modelled using compartments from an SEIR model, with Susceptible, Exposed, Infected, and Removed individuals. The incubation period for the disease is assumed to be 7 days, and the infectious period to be 21 days (following, for example, Refs [9] and [14]). Sojourn times in each of these compartments are assumed to follow an exponential distribution.

Agents can transmit the disease amongst each other in one of two channels: either sexually (for MSMs) or non-sexually (for everyone). We assume that the greatest transmission of the disease between agents via non-sexual contacts occurs at home. The parameter $\beta$ controls the force of infection at the home. This parameter is varied, to see how the spread of the disease varies with the disease transmission strength. In addition, the model also allows for a weak probability of transmission in the workplace as well, assuming a reduced level of physical contact leading to infection.

In addition to individuals moving between homes and workplaces in 12 hour schedules, MSM agents in our simulations can also meet other agents in their sexual contact network through a sexual "encounter". This is modelled in the BharatSim framework as a "`behaviour`" (see Section 2.2 of the main text). The probability distribution of the interval between which an individual MSM meets a contact in his network is assumed to be an exponential distribution with a mean of 7 days.

During such a sexual encounter, if either of the two MSMs is infected, they can infect the other, with some probability $\mu$. In the absence of empirical data on this parameter, we vary $\mu$ from 0–100% in our simulations, as done in Ref [9]. Other modelling studies (see Ref [14], for example) assume a single value of 20%. We note that $\mu$ can also be used to model the usage of prophylactic measures like condoms, which have been found to reduce the risk of mpox spread [15].

#### 9.4 Results

We simulate the spread of the disease by assuming an initial infection seed of a single MSM in the population. In Fig S9.2 we show the total infection curves for different values of the transmission parameter $\beta$ and the probability of infection per sexual encounter $\mu$. The different curves in each of the panels represent different values of $\mu$. The figures show the average results over 100 simulations, but it is worth noting that there is substantial variance, with a good portion of runs dying out without percolating to the entire network.

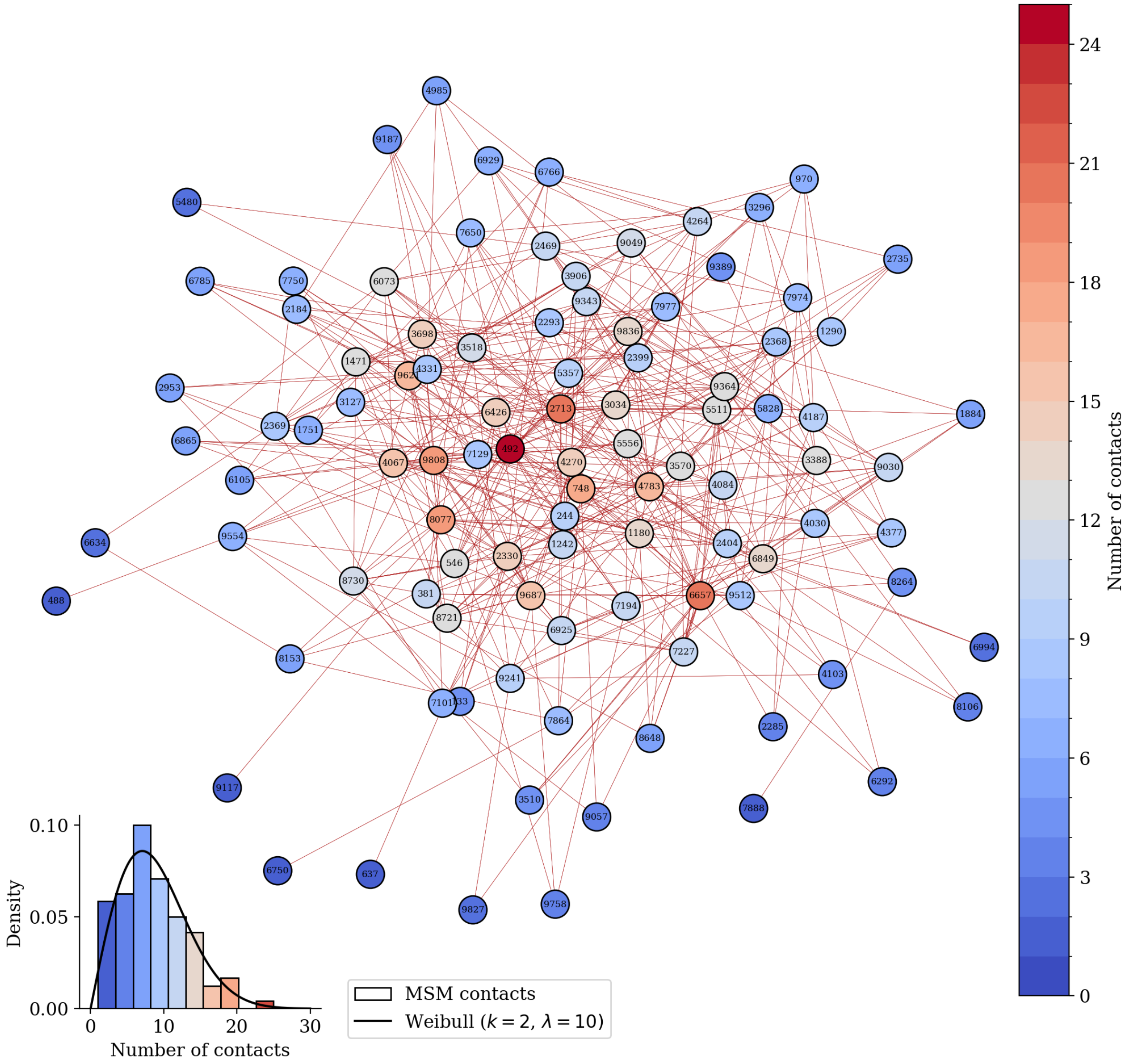


**Fig S9.1:** The contacts of the MSMs in the population as assumed to be distributed as a Weibull distribution. The statistics of the distribution were chosen such that they fall within the range of the results shown in [12]. In the inset we show the actual distribution of contacts, compared with a fitted Weibull distribution with parameters $k = 2, \lambda = 10$.

We then explore the possibility of getting similar results by starting with an initial infection seed involving non-MSM agents. We find that in order to get an outbreak size that is comparable, we require a much higher initial infection seed; this has also been seen in related work [9]. In Fig S9.3, we run a similar set of simulations which we seed with 50 non-MSM agents. Interestingly, the initial spread of the disease is dominated by the non-MSM population. During this spread there is no dependence on $\mu$. However, once the disease enters the MSM population, a strong dependence on $\mu$ is seen, and a second epidemic peak can be observed, whose dynamics is driven by the MSM population.

In addition, if we now include a very small probability of infection at the workplace (1% the rate of infection in the household), these effects are further amplified, as shown in Fig S9.4.

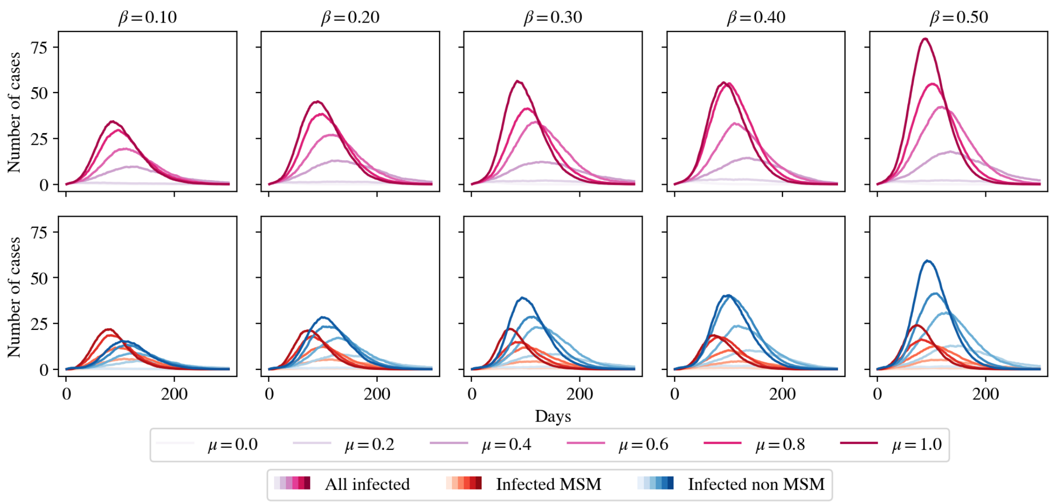


**Fig S9.2:** Infection curves for different values of household infectivity $\beta$ and probability of sexual transmission $\mu$, when the infection is seeded with 1 MSM agent. The top row of panels shows the the total number of active infections in the population in each scenario, while the bottom row shows the division of cases between the MSM and non-MSM population. The red curves show the spread in the MSM population, while the blue curves show the spread in the general population. Darker colors indicate higher values of $\mu$. We see that the peak in the general population is shifted further to the right as compared to the MSM peak. Additionally, we find that the MSM peak is more or less independent of the parameter $\beta$, when compared to the general population peak, however both of these peaks depend strongly on the parameter $\mu$. The curves are averages over 100 runs.

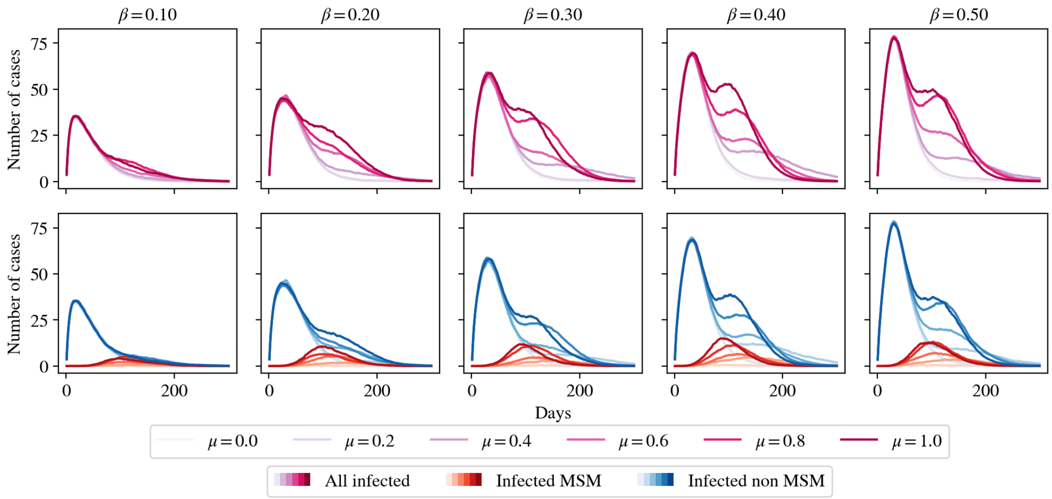


**Fig S9.3:** The same results as in Fig S9.2, but with an initial infection seed of 50 non-MSM agents. We find that this higher initial infection seed is needed to get comparable outbreak sizes. While the initial spread of the disease is amongst the non-MSM population and therefore independent of $\mu$, we see that a secondary peak is observed once the disease enters the MSM network, post which a strong dependence on $\mu$ is found. Again, the curves are averages over 100 runs.

#### 9.5 Discussion

Our results illustrate how the movement of the infection between MSM and non-MSM networks can lead to complex structure in the progress of infection, involving at least two time-scales. Notably,

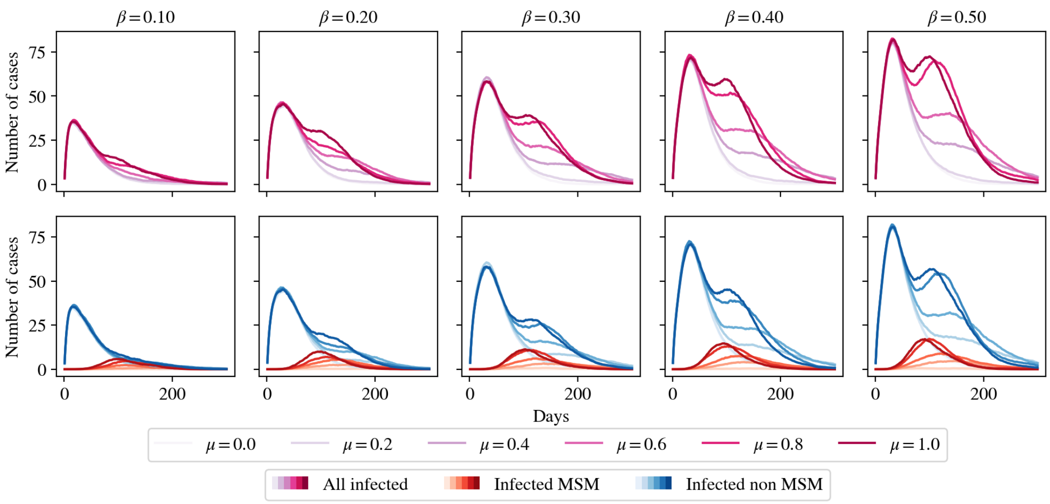


**Fig S9.4:** Continuing the assume an initial infection seed of 50 non-MSM agents, we find that if we include a small probability of being infected in the workplace (1% the probability of household infection), the results in Fig S9.3 are further amplified. The curves are averages over 100 runs.

the spread initiated in the non-MSM population can produce an initial peak that can then give rise to a second peak once it enters the MSM network. We also illustrate how a long tail of infection can be produced as a consequence of even weak spread involving transmission in the workplace. This has clear implications for control.

The results here may be useful to public health policy as it pertains to mpox control in the Indian context, especially since the effectiveness of prophylactic measures such as vaccinations, applied to different sections of the population, can be assessed using BharatSim.
